# An automated Dashboard to improve laboratory COVID-19 diagnostics management

**DOI:** 10.1101/2021.03.20.21253624

**Authors:** Emma Maury, Marc-Olivier Boldi, Gilbert Greub, Valérie Chavez, Katia Jaton, Onya Opota

**Affiliations:** Faculty of Business and Economics, University of Lausanne, Switzerland; Institute of Microbiology, Lausanne University and University Hospital of Lausanne, Lausanne, Switzerland; Infectious Diseases Service, Lausanne University and University Hospital of Lausanne, Lausanne, Switzerland

**Keywords:** CoVID-19, SARS-CoV-2 RT-PCR, microbiology diagnostic, dashboard, automation, digitalization, reliability, laboratory management, quality management

## Abstract

**Background:** In response to the CoVID-19 pandemic, our microbial diagnostic laboratory located in a university hospital has implemented several distinct SARS-CoV-2 RT-PCR systems in a very short time. Thanks to our automated molecular diagnostic platform, more than 140’000 SARS-CoV-2 RT-PCR tests were achieved over 12 months, with peaks higher than 1’500 daily tests. A dashboard was developed to give access to Key Performance Indicators (KPIs) to improve laboratory operational management.

**Methods:** RT-PCR data extraction of four respiratory viruses – SARS-CoV-2, influenza A and B and RSV – from our laboratory information system (LIS), was automated. Important KPIs were identified and the visualization was achieved using an in-house dashboard based on the open-source language R (Shiny). Information is updated every 4 hours.

**Results:** The dashboard is organized into three main parts. The “Filter” page presents all the KPIs, divided into five sections: i) general and gender-related indicators, ii) number of tests and positivity rate, iii) cycle threshold and viral load, iv) test durations, and v) not valid results. Filtering allows to select a given period, a dedicated instrument, a given specimen, or a requester for instance. The “Comparison” page allows a custom charting of all the available variables, which represents more than 182 combinations. The “Data” page gives the user access to the raw data in table format, with the possibility of filtering, allowing for a deeper analysis and data download in Excel format.

**Conclusions:** The dashboard, that gives a rapid access to a huge number of up-to-date information, represents a reliable and user-friendly tool improving the decision-making process, resource planning and quality management. The dashboard represent an added value for diagnosric laboratories during a pandemic, where rapid and efficient adaptation is mandatory.

## 1 Introduction

In December 2019, a new virus causing pneumonia of unknown etiology has emerged in China. Its incidence exploded very quickly, first in the Wuhan region (Hubei province), then in the other regions of China and other countries in Southeast Asia. On January 30, the World Organization of Health (WHO) declared this new coronavirus as a “public health emergency of international concern” [1]. On the 20^th^ of February 2020, a first patient was diagnosed in Italy in the Lombardy region. The epidemic has since spread to other European countries, including Switzerland [2] and a first case was admitted in our hospital already on 28^th^ February. On the 11^th^ of March, 2020 that WHO declared Pandemic Coronavirus Disease 2019 (COVID-19) [3-5].

To face the COVID-19 pandemic caused by the severe acute respiratory syndrome Coronavirus 2 (SARS-CoV-2), diagnostic laboratories had to develop reverse transcriptase polymerase chain reactions (RT-PCR) tests allowing the detection of SARS-CoV-2 RNA in patients with suspected COVID-19. Our laboratory, the Institute of Microbiology (IMU), located in one of the five teaching hospitals of Switzerland, the Lausanne University Hospital (CHUV), rapidly developed RT-PCR to detect SARS-CoV-2 in clinical specimens [6]. Microbiological diagnosis of SARS-CoV-2 represents one of the pillars of the diagnosis of COVID-19. Indeed, RT-PCR is also the heart of the patient care and epidemic control process and will be the mainstay of several clinical studies. Although our laboratory has extensive experience in the development of RT-PCR, the introduction of this new parameter represented a challenge in terms of speed of development [7]. It is also the first time that an introduced parameter has been used on such a large scale in such a short time; more than 10’000 tests were carried out in one month in Spring [6] and even in a single week during fall 2020. This was possible thanks to automation and digitalization, to allow high throughput and acceptable time to results [7]. In this context, the IMU set strategies to ensure the quality and reliability of the RT-PCR. This included the monitoring of key performance indicators (KPIs) for quality management such as the proportions of positive tests or the virus load, both per day, per instruments, and per requester. These indicators aimed for instance to identify variations not explained by epidemiological changes. Indeed, abnormal variations could be synonymous with pre-analytical problems (sampling problem, transport medium, etc.) or even analytical problems (mutation in the target sequences of PCRs associated with losses of sensitivity or specificities). The IMU also defined KPIs for operations management such as the time to results [8].

Before COVID-19, such indicators were monitored periodically, for example in the context of an annual report or retrospective studies. At the beginning of the COVID-19 outbreak, the IMU decided to follow these indicators frequently. Because the needed manual analyses were time-consuming, the monitoring of analytical and operational KPIs was carried out once a week initially and then, twice a week depending on the period. These analyses were also prone to error, due to multiple sources of information, repeated manual actions (e.g., copy/cut and paste) and the diversity of the data.

A dashboard is a graphical user interfaces (GUI) with a data base. It allows to retrieve the relevant information, often KPIs, in a specific context by representing the data in a meaningful and usable way [9]. For a detailed presentation, see [10]. In the management and business contexts, dashboards aim at reversing the overwhelming information volume into an opportunity [11] and are part of visual analytics, defined as the “science of analytical reasoning facilitated by interactive visual interface” by Cook and Thomas [12].

Like any other information technology in the healthcare industry, which affects positively its efficiency [13], dashboards help monitor daily activities [14], such as tracking ongoing operations, a priority in healthcare institutions [15]. Providing an easy access to this information helps the team to make better informed decisions [16], which could take a tremendous amount of time without the technology [17]. Correctly designed and built, dashboards improve the institution’s efficiency while providing a better care quality [18]. See Cheng, et al. [19], for an example of a previous study on how to build dashboard to track respiratory viruses, such as Influenza.

With COVID-19 spreading fast over the world, the speed at which data are gathered, integrated, and used became central in the management of this crisis by all health-involved institution teams. Interactive dashboards appeared to be appropriate to this aim. A famous example remains the one from John Hopkins University [20]. In Switzerland, corona-data.ch [21] is an up-to-date webpage built at the macro level. The whole healthcare industry being impacted by the pandemic, various topics and areas were analyzed through dashboards: e-consultations [22], incident command [23], performance comparisons to similar institutions [24].

Also, laboratories responsible for testing patients during an outbreak must control some information to ensure the highest quality of results. In particular, it is crucial to define KPIs, for example to better track the daily operations [25] ensuring enough testing capacity. Moreover, providing valuable insights and pieces of information to the laboratory management can be critical when the objective is to increase capacity and quality as well as maintaining schedules [26].

A dashboard is an expression of a database. Therefore, like mentioned by O’Donnell and David [27], the resulting decision process depends on the Information System (IS), the environment, and the user skills. Regarding the dashboard content, there is no consensus on the format of visualizations: from no effect on the user’s judgement [28] to the lack of universality in representation [29], or a preference for tabular information [30]. Wilson and Zigurs [31] showed that even the user’s preferred format did not lead to greater performance, except for symbolic and spatial tasks [32]. Choosing the appropriate visualization can be challenging and is subject to various principles. Lengler and Eppler [33] condensed many visualizations into a Periodic Table of Visualization Methods, according to which the dashboard content can be classified into three categories: boxes, tables and plots. Each has advantages and drawbacks, the choice being made based on the end-users needs.

In this paper, we present the design, the development, and the use of a dashboard targeted to a laboratory located in a teaching hospital, in charge of PCR test, following the COVID-19 outbreak, such as the IMU. This includes i) to define the need, ii) to build the dashboard, iii) to deploy the tool, and iv) to demonstrate the added value in terms of the quality and operations management for the laboratory mission. This research focuses on aspects other than epidemiological matters (patient type, pathogen, period of the year, etc.), and which can explain some variation of results in the laboratory. We split these aspects into two main categories: quality issues and management issues.

## 2. Material and Methods

### 2.1 RT-PCR and Data

RT-PCR for the detection of SARS-CoV-2 from clinical specimen where achieved as previously described using our in-house molecular diagnostic platform (MDx platform), the Cobas SARS-CoV-2 test on the Cobas 6800 instrument (Roche, Basel, Switzerland), and the Xpert Xpress SARS-CoV-2 assay (Cepheid, Ca, USA) [6,34,35]. Viral load were obtained by conversion of the Ct (Cycle threshold) values of the instruments using either a plasmid containing the target sequence of the PCR obtained from RD-Biotech (Besançon, France) or using purified viral RNA, kindly provided by the Institute of Virology of the University of Berlin, la Charite [34,35]. The dataset feeding the dashboard is an extract from Molis, an IS used at the IMU. The extract is performed every four hours. The analyses of four respiratory virus of interest were flagged in the system: SARS CoV-2, Influenza A and B, and the Respiratory Syncytial Virus (RSV). A comma-separated values file (csv) with new observations validated in the past four hours is uploaded in a specific folder to be read through the dashboard.

To date, the system has more than 140’000 observations, of which the SARS-CoV-2 accounts for more than 96%. For each sample (swab, blood, etc.), the available entries include a unique ID, date of birth and gender of the patient, its hospitalization status, an anonymized code of the entity requesting the test (doctors, clinics, other laboratories, etc.), the date-time of the sampling (when available), of the test, and of the result sending as well as the type of sampling (nasopharyngeal or oropharyngeal secretions, blood sample, etc.). The original dataset also contains analysis codes showing the test result (positive, negative, cancelled, *NOT VALID*), the Ct values, the viral quantification (in copy per milliliter, cp/mL), and whether the analysis had to be repeated.

A sample is related to one patient but that a patient may be tested several times. The analysis codes correspond to a test, which is performed for a specific virus, on a specific device (machine used to perform the test), for a targeted gene.

Some cleaning and data wrangling were performed before building the dashboard. Using a matching table shared by the IMU, the analysis codes were renamed using more user-friendly structure (NOM.VIRUS_TYPE.ANALYSE_APPAREIL_GENE). Then, different measures are extracted, especially on date-time data: the reception duration is the difference between the sampling time and the reception time at the laboratory, the test duration is the difference between the reception time and the results validation time and the total duration is the sum of the last two. Using the date of birth, patients were categorized into age groups with a 10-year window. Similarly, the type of sampling was recoded using wider groups categories, the most present being NPS (nasopharyngeal secretions). Then, each analysis is assigned a “Virus” and a “Device”, corresponding to the non-empty analysis codes described above. Another “Confirmation” variable is added, showing whether the analysis had to be reperformed.

Finally, four different tables are created, corresponding to each virus present in the dashboard.

### 2.2 Platform

The interactive dashboard was built using Shiny [36], based on the opensource programming language R [37] in the RStudio Interactive Development Environment [38]. CSS and JavaScript were also used to tailor the dashboard to the needs of the end-users. The dashboard relies on several packages: *shinydashboard* [39] for the page structure, *plotly* [40], and *ggplot2* [41] for interactive graphs, *DT* [42] for interactive tables, *shinyjs* [43] for some custom interaction, *shinyWidgets* [44] for input objects and *readxl* [45], *plyr* [46], *dplyr* [47], *lubridate* [48], *tidyr* [49], *tidyverse* [50], *stringr* [51], *psych* [52] and *forcats* [53] for data wrangling. The dashboard runs locally on every user’s computer, in order to prevent from security issues.

The dashboard is built in two dimensions (Figure 1). The horizontal dimension is the *Target* and the vertical dimension, the *Action*. The level of detail and the amount of possibilities increase when the user goes down on the *Action* sections.

**Figure 1:**
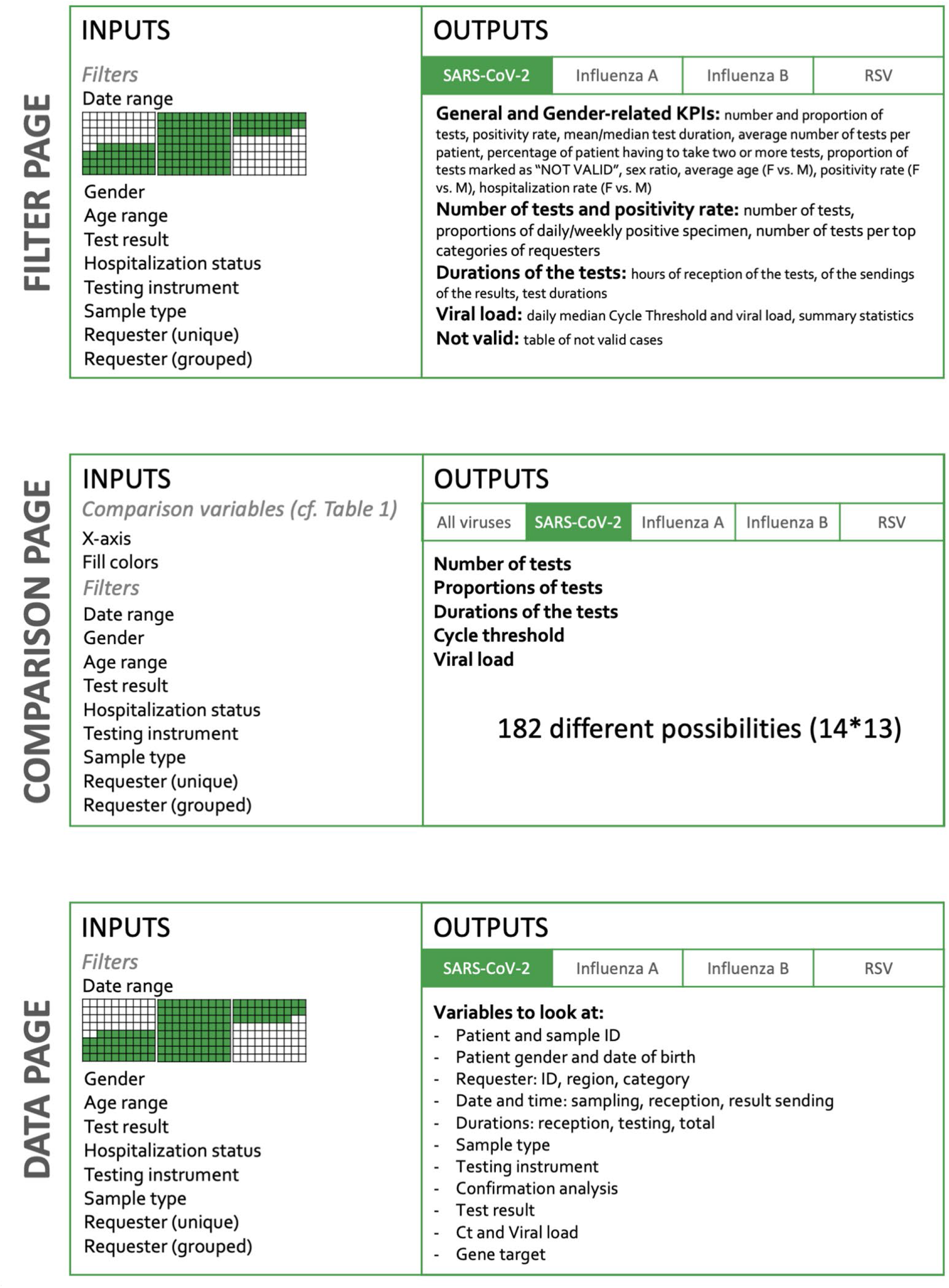
Dashboard Structure. The dashboard is organized in three main parts: *Filter* page, *Comparison* page and *Data* page. On the filtering page, key indicators are available such as the number of samples, time to results, positivity rate, percent of invalid tests, etc. These indicators are provided by default for the whole dataset, but are also available for subgroups, according to the filtering criteria applied to the whole dataset. Thus, it is possible in a click to observe specifically the tests done during a given period or done on a dedicated instrument. It is also possible to select only the analysis performed for a given requester. The *Comparison* page offers more than 182 combinations of the KPI.The *Data* page, gives access to the raw data in a table format that can be downloaded; filters can be applied to shoose a subset of data of interest.

As stated above, picking the appropriate visualization can be challenging and is subject to various principles. The main components chosen to build the dashboard are: 1) infoboxes: this type of visualizations gives an immediate information on some key metrics. Placed at the top of the dashboard, the user directly sees crucial information. When having several of these, it is important to group and label them appropriately [54], 2) tables: columns describe a specific attribute for each row, showing the user a detailed view, ready for a deeper inspection. Filtering and ordering options are available to display the part of interest of the data and 3) plots: we used both Abela [55] and the data-to-viz.com tool created by Holtz [56], which provide clear tools to select the appropriate chart relative to the data. This dashboard mostly uses Column charts and Stacked Column charts, Scatter plots, Line charts and Boxplots. The latter has the advantage of displaying many information at once, and the end-users are accustomed to this format.

Finally, for a quicker adoption and an optimal usage, the dashboard was built in French, the mother tongue of the end-users.

### 2.3 Ethics

The data were obtained during a quality enhancement project at our institution. According to national law, the performance and publishing the results of such a project can be done without asking the permission of the competent research ethics committee.

## 3. Results

### 3.1 Structure and main KPIs of the dashboard

#### Global structure of the dashboard

The content of the dashboard was guided by three main principles: comprehensive initial brief, multiple feedback loops, and close monitoring of latest breakthrough discoveries about Sars-CoV-2. At the beginning of the process, the end-users, namely the managers of the laboratory of molecular diagnostic, formulated their main needs and made suggestions on what to report in the dashboard. This included the main KPIs, such as the number of tests per day as well as some of the inputs that could be entered by the user, for instance filters such as RT-PCR instruments or patients’ gender or age. In an AGILE-like methodology, applying continuous improvement [57], demos were regularly performed and feedbacks shortly implemented. The SARS-CoV-2 being a novel virus, the scientific literature was closely scrutinized to incorporate new relevant elements, such as gender-related indicators, which were included after [58] published their research on gender bias in Covid-19 diagnosis.

The current version of the dashboard contains three main pages detailed in figure 1: i) “*Filter”*, ii) *“Comparison”* and iii) *“Data”*. The *Filter* page allows the user to select inputs such as a date range, the gender, the age, the test result, the hospitalization status, the device used, the confirmation status, the type of sample, and the type of requesters. In the *Comparison* page, the user selects variables to appear on graphs, but also filters the dataset to narrow down comparison subjects. Finally, the *Data* page also lets the user filter the dataset to look at individual observations. Overall, the user therefore has a role in filtering observations and in deciding which information to be represented (Figure 1 and suppl. Table 1).

**Table 1.**
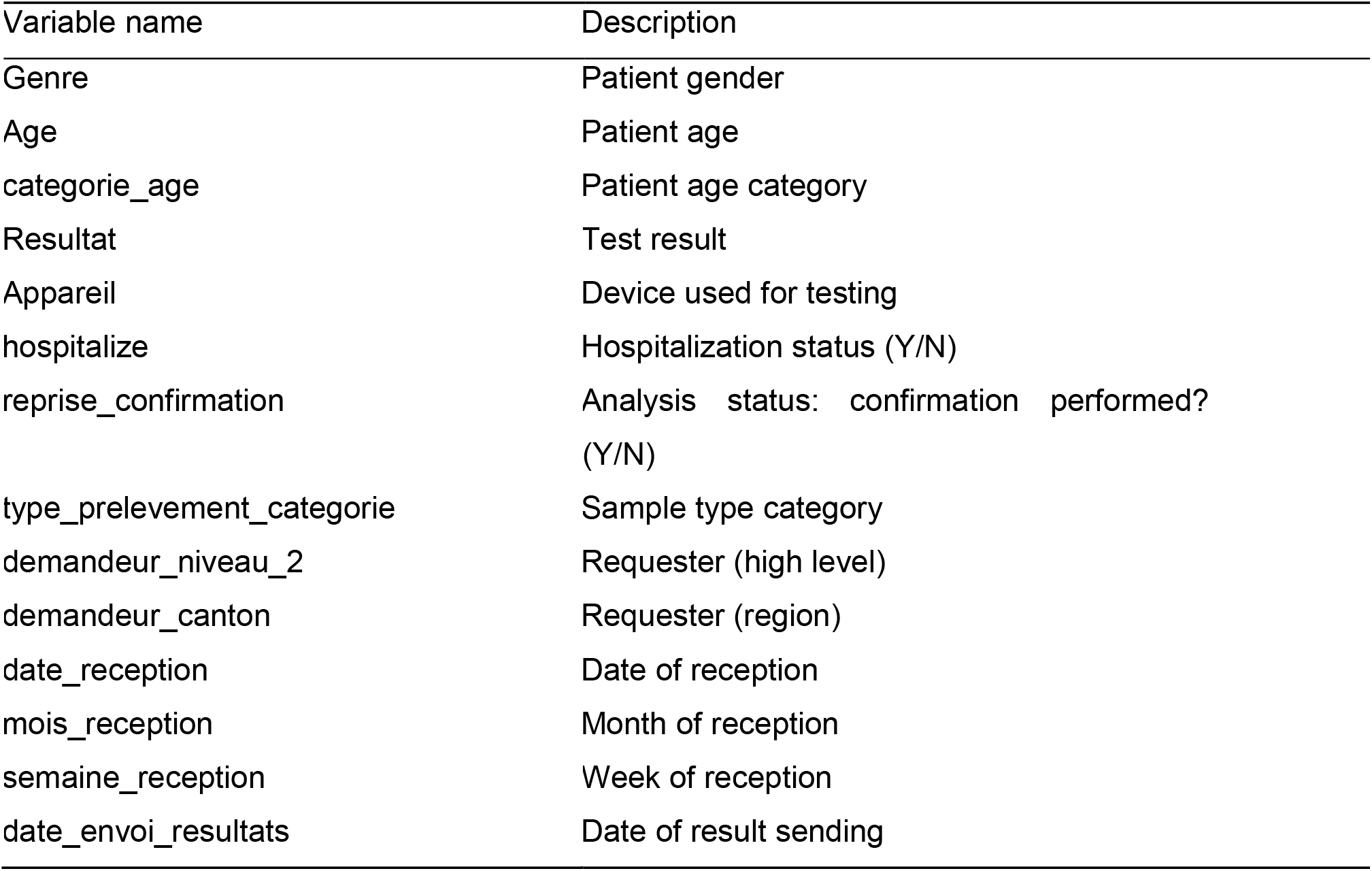
available variables for custom charting in the Comparison page.

#### The Filter page

The Filter page presents a total of 10 KPIs and is divided into five sections: general and gender-related indicators, number of tests and positivity rate, viral load, test durations, and “NOT VALID”.

##### General and gender-related indicators

The ten KPIs are presented in aggregate form in this section, some of them being detailed in the subsequent sections. The most general figures are shown at the top of the layout. This first section is split into two groups. The “General Indicators” groups i) three workload-related KPIs: the number of tests performed for the selected virus, the proportion of the tests after filtering, the number of positive tests, the positivity rate, the average test duration displayed next to the median duration, ii) three quality related KPIs: the average number of tests per patient, the percentage of patient having to take two or more tests, the proportion of tests marked as “NOT VALID” and iii) gender-related Indicators including the sex ratio (positive women out of positive men), the average age, the positivity rate and the hospitalization rate (Suppl. Figure 1.A and 1.B).

##### Number of tests and positivity rate

In this section, the number of tests and the proportions of daily positive specimen are detailed. The number of tests is presented per day of reception at the laboratory, per date of sending the results, and per age category. The proportions are presented per day and week of reception. In addition, an interactive table shows the number of tests per top categories of requesters. The plots are interactive to avoid cluttered visualization while allowing the user to choose the appropriate representation (zoom, labels, etc.) (Figure 2.A and 2.B).

**Figure 2.**
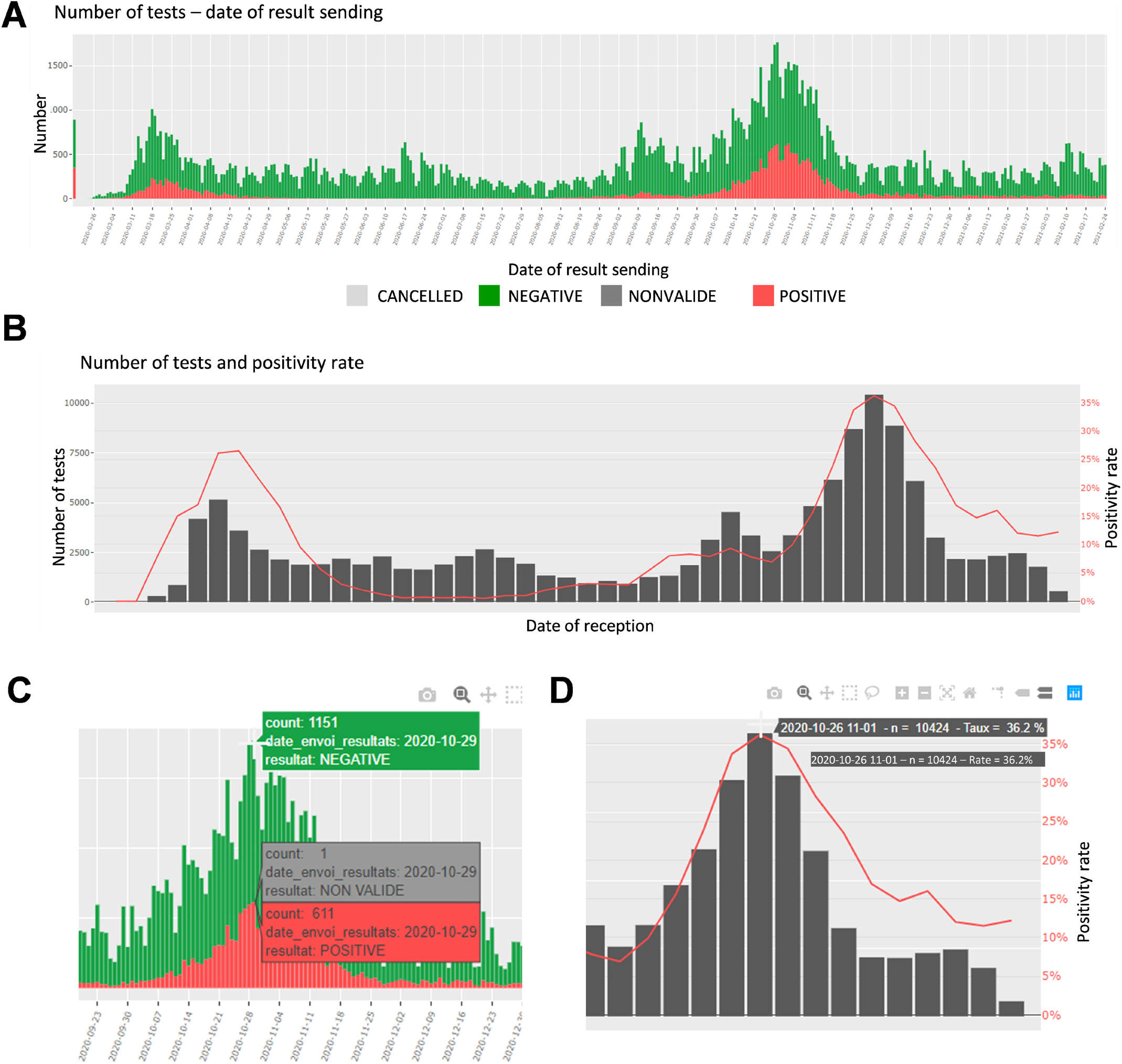
Screen shots of the upper panel of the Filter page. **A**. Number of tests and test results per date of results sending. **B**. Weekly number of tests and positivity rate. **C**. Panel A zoomed on the label shown when hovering. **D**. Panel B zoomed on the label shown when hovering.

##### Durations of the tests

In this section, the user finds the hours of reception of the tests and of the sending of the results, along with the test durations. These are presented by test results (positive or negative); the number of tests is displayed for each hour of a 24-hour day (Figure 3.B). The average test durations and the average reception durations are displayed below for each day of sending, respectively day of reception.

**Figure 3.**
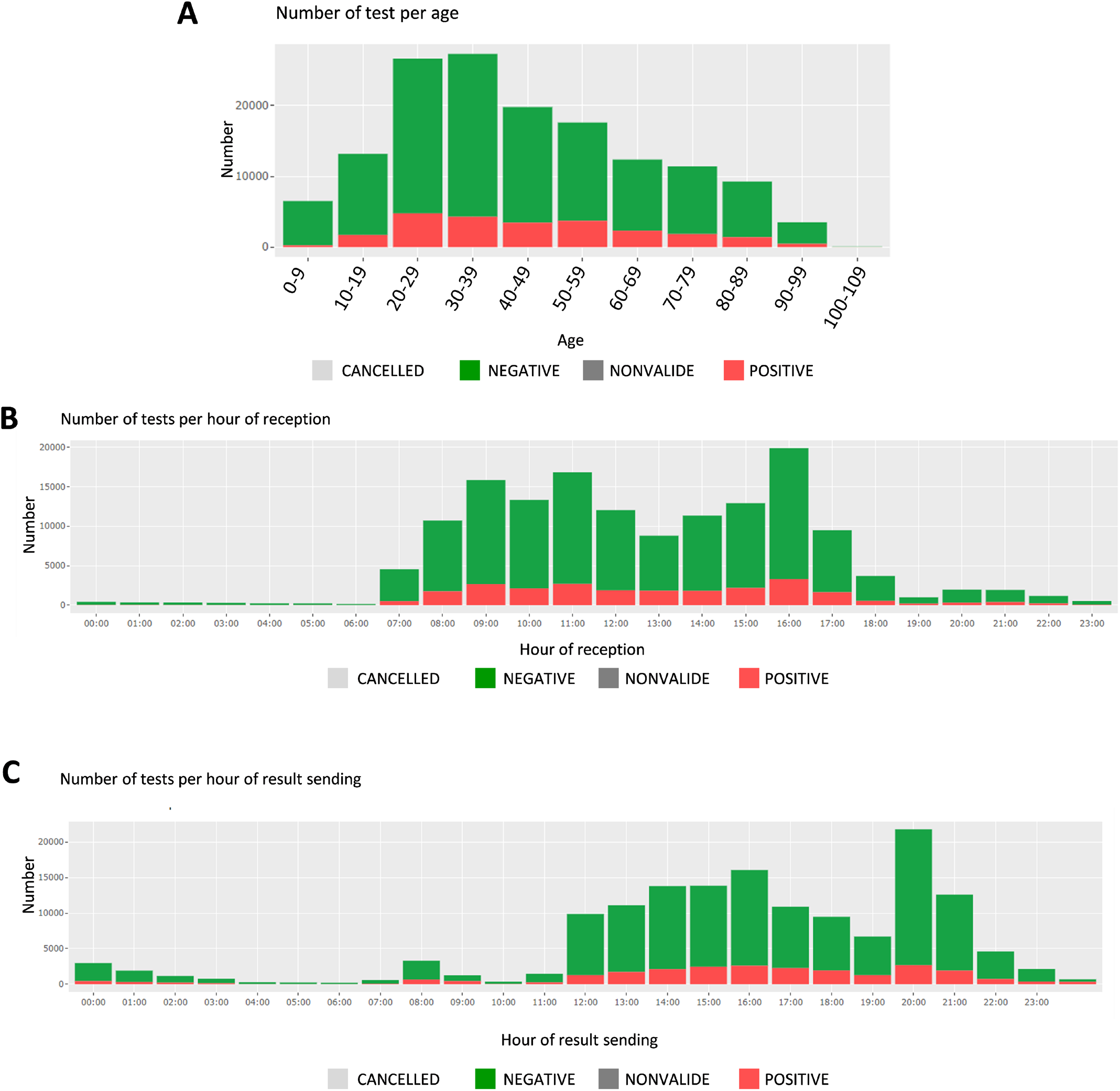
Screen shots of the middle panel of Filter page showing. **A**. The number of tests and test results, per age group, **B**. The number of tests per hour of reception and **C**. The number of tests per hour of results sending.

##### Viral load

This section focuses on the viral load and Cycle Threshold (Ct), a KPI not presented in the first section. First, the median Ct for each day is showed, grouped by analysis (each device, each gene, each repetition); in total 12 analyses (Figure 5). The viral load is also available in copy/millimeter (cp/mL) since April 9th, 2020, date from which the laboratory started to keep this record. These are shown in a scatterplot crossed by type of sampling (blood, in nasopharyngeal secretions, etc.) as well as in a summary statistics table (Suppl. Figure 3).

##### Not valid

Finally, a descriptive table of the cases for which the result of the analysis was “NOT VALID” is displayed. The user can then identify any issue and investigate further. For sake of readability only some variables are shown (Suppl. Figure 4).

#### Comparison page

When moving to the *Comparison* page, the user sees first a global summary for all viruses. It shows the daily number of tests for each disease as well as the corresponding daily positivity rate. Then, like the *Filter* page, each virus information is shown in a specific tab. For each virus, the user can select the *x*-axis variable and the coloring variable. An error message is displayed when the chosen variables are the same (Table 1).

Upon this choice, the absolute and the relative number of tests are plotted, such as shown in Suppl. Figure 5 for gender and week of reception. The following plot shows data about the test duration. An example in Figure 4.B shows the average test duration per instrument, depending on the result of the test. This is especially useful to control the speed of some devices. Finally, the Ct information is displayed using boxplots (Figure 6.A), a familiar representation to the end-users. All the described figures are common to all viruses. For SARS-Cov-2, an additional boxplot of the viral load in log_10_ scale is displayed (Figure 6.B). This additional boxplot appears only for this virus whose load can vary from 1000 to 14000000000 cp/mL. The log_10_ scale provides a more readable graph.

**Figure 4.**
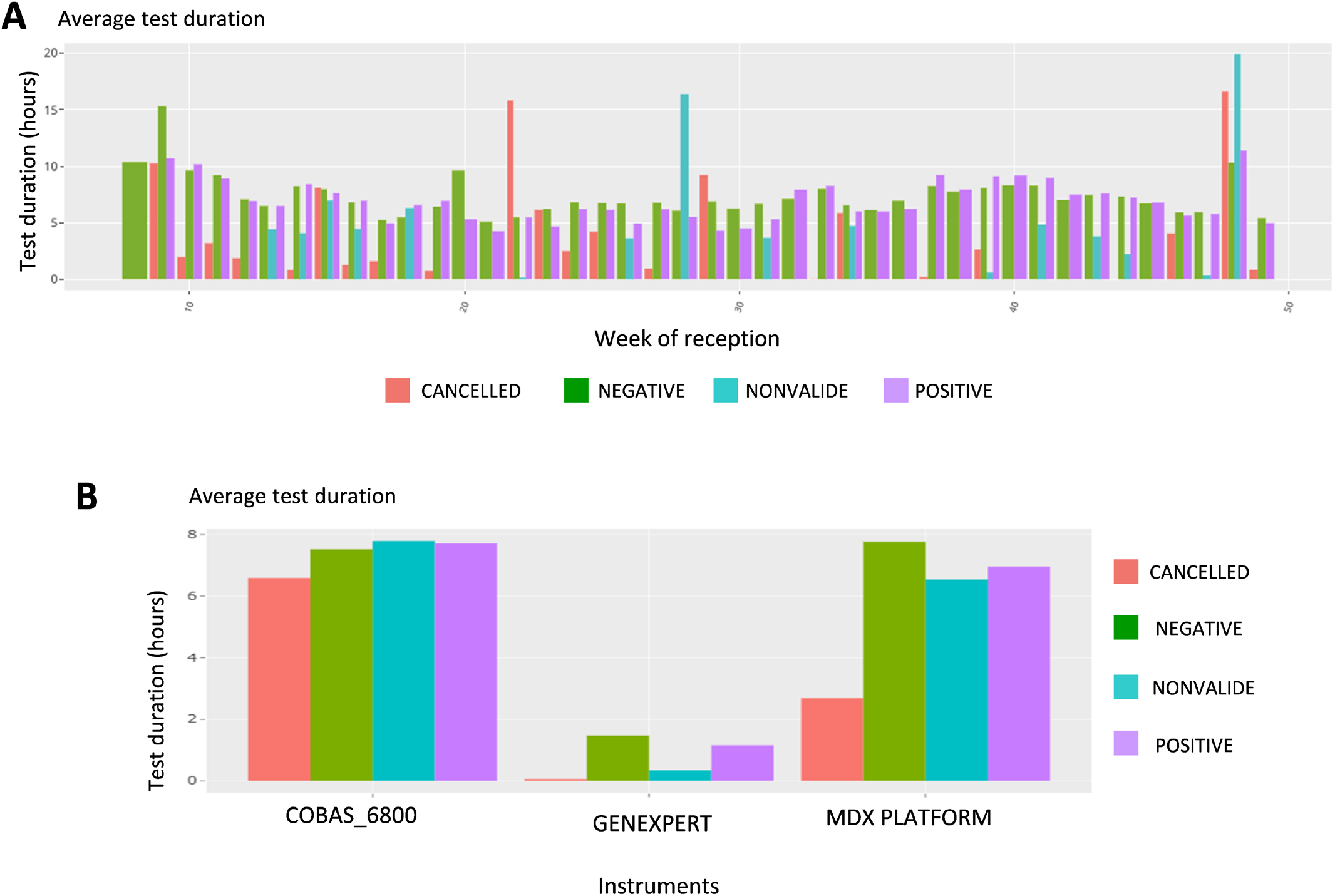
Screen shot of the Comparison page that allows 182 different combinations; focus on tests duration. **A**. Average test duration, per week of reception and test result. **B**. Average test duration, per device and test result.

**Figure 5.**
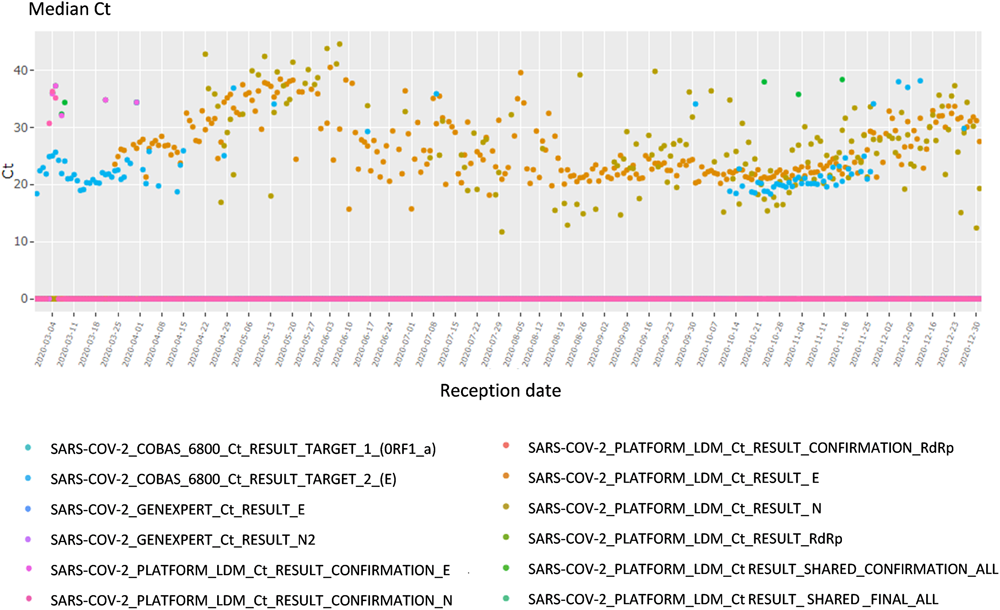
Screenshots of the lower panel of the Filter page. Cycle threshold time series plot, per date of reception.

**Figure 6.**
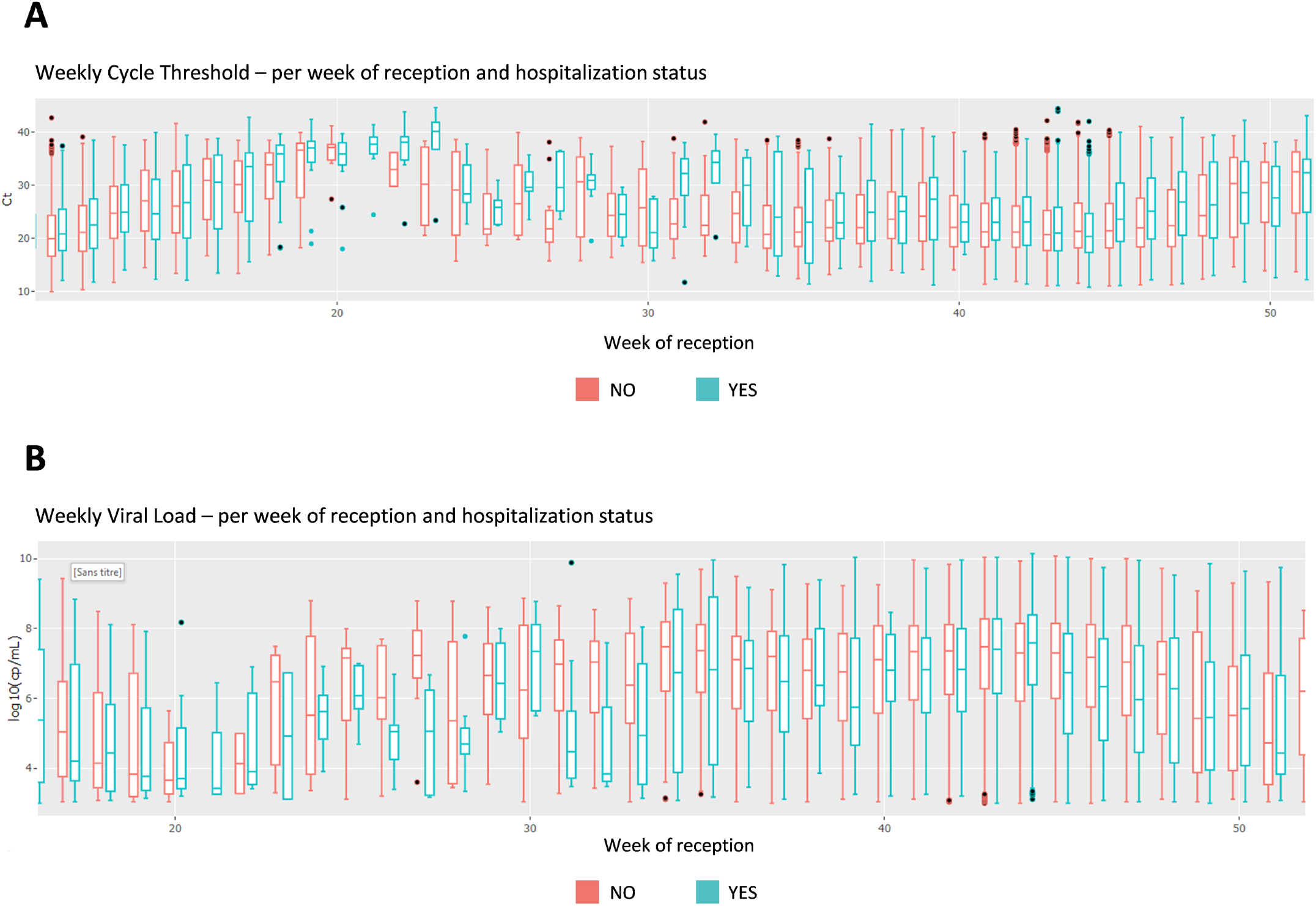
Screen shot of the Comparison page that allows 182 different combinations, focus on cycle thresholds (Ct). **A**. Cycle Threshold boxplots per hospitalisation status, per week of reception. B. Comparison page - Viral load boxplots per hospitalisation status, per week of reception

#### Data page

On the *Data* page, the users can have a look at the raw data in a table format, after filtering if wanted, thus deepening the analysis of an issue located with the previous pages tools. They can then download the data and explore it in Excel. This simple feature appeared to be surprisingly useful to the users who can then extract data more quickly than with a data base request from Molis.

### 3.2 Operational management improvement

The general KPIs (e.g., number of daily tests, average number of tests per patient, etc.), such as Suppl. Figure 2.A, appearing at the very top of the dashboard are used to continuously reassess the current situation. Seeing the evolution of the past few days or weeks helps the staff adjust their resources: launch some recruitment or send back the technicians from other department, adjust the repartition of staff among the laboratory, handle material shortage by anticipating future volumes, etc.

On a regular basis, the KPIs regarding the durations of the test allow the detection of operational issues. For instance, the dashboard led to the discovery of a huge amount of samples received at the end of the business day, and helped tune the process of deliveries at the laboratory. This is central to meet the needs of the hospital, of the requesters and, at the end, of the patients themselves. The tool also supports the team in charge of identifying potential causes of delay: by being able to look into various combination and level of details, it is possible to point out these causes during the whole process (from sampling, to testing, to result sending). They hence can then be investigated individually (e.g., by requester) or more globally (e.g., by day of the week). Figures 3.B and 3.C are used on a daily basis to continuously reassess this crucial aspect of timing.

The dashboard also comforted some intuitions the institute had from its experience: it helped identify the peak hours, adapt the distribution of the employees, and make recommendation to major requesters in order to smooth the operational activity during the day.

Finally, the daily tracking of these KPIs allowed the inventory monitoring of materials and avoided an unexpected shortage of scarce test resources.

### 3.3 Quality management improvement

#### Positivity rate

Having the possibility to observe the positivity rate by instrument can help diagnose any failure, such as contamination. For instance, an increase in the positivity rate could be due to a contamination of the test reagent. Easily accessing this information, (Suppl. Figure 2.B for an example) being able to quickly analyze it and linking it to Ct values provides a critical advantage in maintaining high quality results over the course of the outbreak. Since the users of the dashboard can also look at the data per requester, or requester type, in case of a sudden variation in the positive rate test results, it allows them to look for non-epidemiologic reasons. Possible explanations include change of sampling methodologies, change of patient type (e.g., from mostly children to elderly patients), addition of new facilities in some requester categories. They can also investigate the geographical origin of the test and share relevant information with the appropriate authorities.

Finally, being able to verify the number of positive specimens by age category is critical, as the patients’ age is playing a key role in the pandemic (spreading role of children, disease evolution among elderly, etc.). Figure 3.A shows the number of tests per age group, which can be filtered for specific periods of time, or different requesters.

#### Viral load

Among various data in the dashboard, the viral load especially helps guarantee the accuracy of the test or conversely identify and solve analytic problems. For instance, any sudden drop or jump in the daily or weekly median viral load raised a warning and calls for explanation (e.g., change of testing strategy, population target). If no explanation is found, one could suspect a problem at the analytical stage (i.e.: pre-analytical stage, virus mutation etc.). A major viral load drop could mean a decrease in analytic sensitivity, which could be dramatic during an outbreak. A decrease in the median viral load was observed in April 2020 which led to a modification of the testing strategy, namely a universal testing strategy [35].

Similarly, tracking the virus loads per patient types helps addressing the test sensitivity. Indeed, since the institute performs tests on a large variety of patients, it needs to track this measure to adjust the tests sensitivity in real time, if relevant.

Additionally, the viral load by type of specimen (nasopharyngeal secretions, blood samples, throat secretions…) helps at addressing the performance of the test in different test of specimens [34,59].

The viral load by instrument gives information on the analytical performance of each methods. Indeed, any drop out or increase could be the result of virus mutation or contamination.

## 4 Conclusion

To respond to the CoVID-19 pandemic, our laboratory had to introduce several methods to detect SARS-CoV-2 in clinical samples by RT-PCR within a short time. In fact, this sensitive method is reliable to control the spread of the virus by identifying infected patients, including asymptomatic subjects [35,60]. As a result, the number of RT-PCR tests performed each day increased rapidly with an average of 350 tests per day with peaks beyond 1000 tests per day during the first epidemic wave in Switzerland and more than 1700 tests per day during the second wave.

Working with the dashboard improved the functioning of the laboratory both in term of management of operations and in term of quality. This improvement was reflected in the reactivity and decision making resulting from the real-time interpretation of indicators. Indeed, this emerging virus posed a large number of challenges for diagnostic laboratories, which had to quickly introduce new diagnostic methods and to adapt to the challenges faced during the different stages of the pandemic.

### Reliability of the RT-PCR tests

The first challenge for diagnostic laboratory was to ensure the reliability of the RT-PCR assays. This was especially important because it is the first time that a test has been used with such a high throughput soon after its introduction. In this, the statistical analyzes proposed by the dashboard help to improve the reliability of these tests. For example, the real-time monitoring of the positivity rate of the tests or of the viral load in the clinical samples enables to highlight analytical problems, which could lead to false positives or false negatives. Using the dashboard, the positivity rate was compared according to the different instruments to check whether one instrument might be associated with a systematic error. However, cautious interpretation of the data available through the dashboard is mandatory since differences might also be due to various other factors. For example, because highly symptomatic subjects admitted at the emergency ward of the hospital were tested with the GenXpert rapid RT-PCR, the tests performed with that instruments were more often positive.

### Management of multiple instruments and stocks

We also had to introduce several different RT-PCR methods. The goal was to guarantee continuity of service in the case of instrument failure or reagent shortage [6], but also to respond to a need for faster tests, notably for analyzes carried out in emergencies to avoid nosocomial infections [35]. In this context of a fleet of several RT-PCR instruments the dashboard allows real-time monitoring of laboratory activity in terms of tests carried out on each instrument. This monitoring has enabled us, for example, to identify a risk of significant penury of the rapid molecular test. This problem was solved by quickly introducing a second test.

#### Time to result

A challenge for the laboratory was to adapt to the needs of clinicians for the patients care and to be useful for public health decisions. This involved a short rendering time. The dashboard allows to dynamically monitor the time of arrival of samples at the laboratory and thus to adjust the work organization. Typically, we added human resources at the end of the afternoon to allow more same-day results.

In its current version, the code is specific to the practice of our laboratory. Thus, it is difficult to generalize to other laboratories or users. So far, the app is still in a development version and suffers from limited performances: slow to refresh, and unlikely to support database growth on the long term.

On the analytical part, there is no multivariate analysis feature. It is thus, for example, not possible to correlate KPIs of several viruses altogether. Furthermore, no automated detection has been implemented yet. And, in the same vein, no efficient forecasting method is available.

We can expect that this type of tool will also help to consolidate the user’s intuitions on the long term. This is linked to the predictive aspect of the dashboard, a topic not addressed in this paper, but a subject of future research. The data accumulation allows to develop models leading to implement (i) automatic alerts (when should resources be increased and when should they be decreased), (ii) predictions (number of positive rates, the number of tests, etc.), (iii) scenario stressing (what would happen if the authorities change the sanitary restrictions?).

These dynamic and automated developments could be based on models for times series forecasting. It may incorporate external factors such as data from other laboratories in the country or other exogenous factors (governmental decisions and meteorological conditions for instance). This could optimize the use of the dashboard for improved impact in the laboratory daily life.

## Data Availability

The data were obtained during a quality enhancement project at our institution and cannot be used for any other purpose

## Acknowledgments

We thank all the staff of the Laboratory of Molecular Diagnostic of the Institute of Microbiology of the University of Lausanne and in particular Miss Zahera Naseri. We also thank Mr Fabien Faverjon, Mr Frank Hottin and the IT and Data Team for their support. All the authors declare no conflict of interest related to this work.

## Supplementary material

**Suppl. Figure 1.**
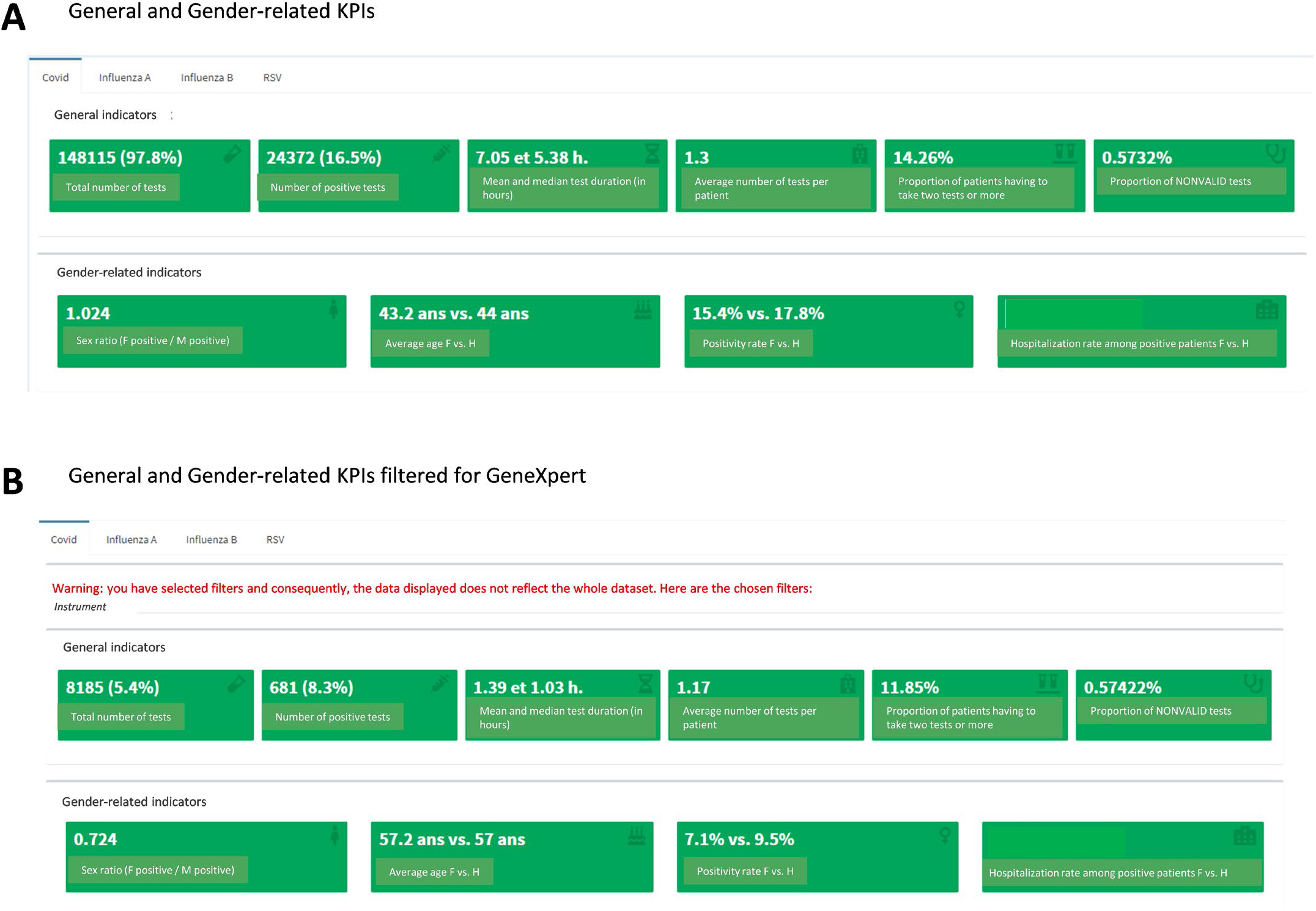
Screen shots of the Filter page. **A**. Number of tests and test results per date of reception. **B**. Positivity rate per date of reception.

**Suppl. Figure 2.**
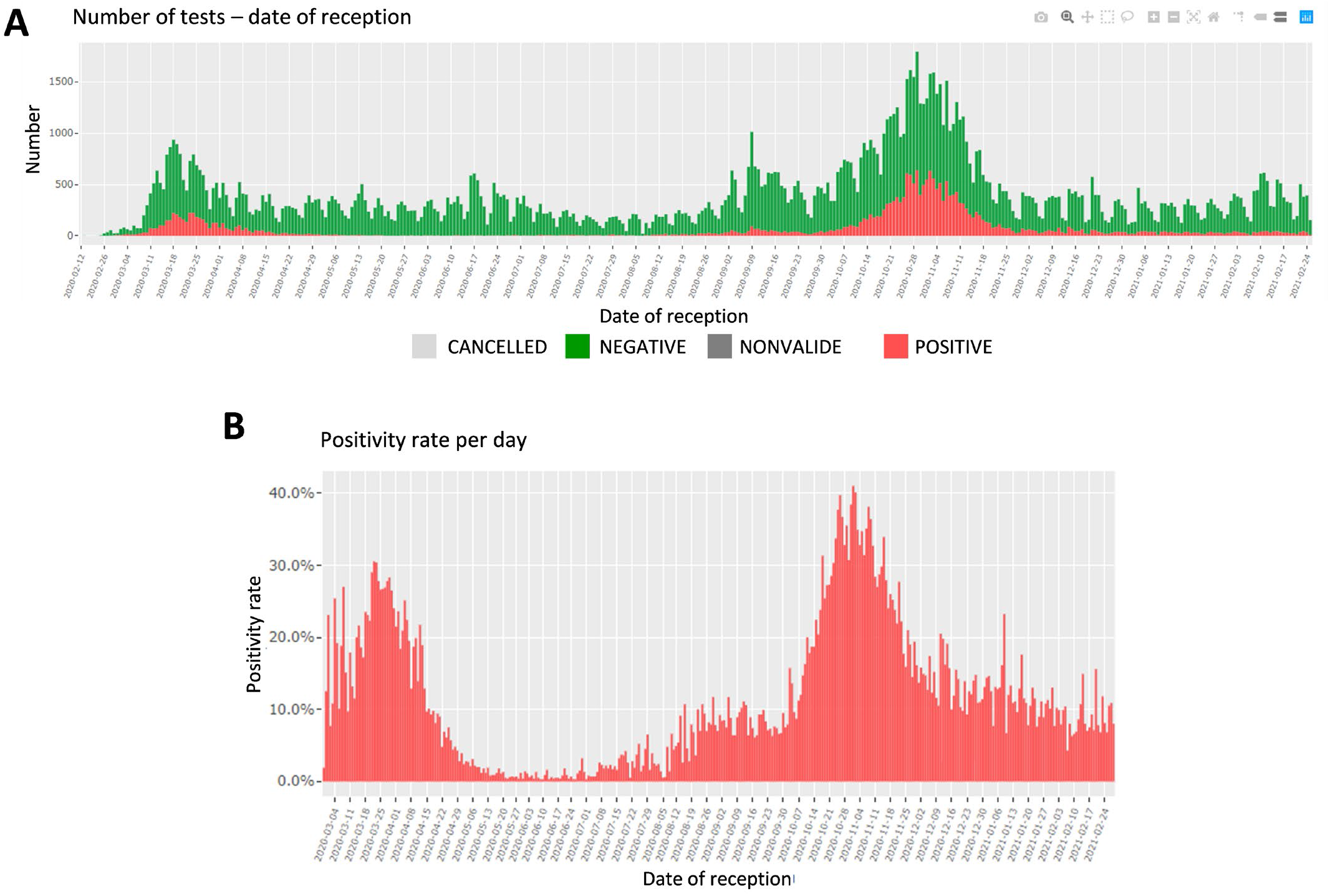
Screen shots of the Filter page. **A**. Number of tests and test results per date of reception. **B**. Positivity rate per date of reception.

**Suppl. Figure 3:**
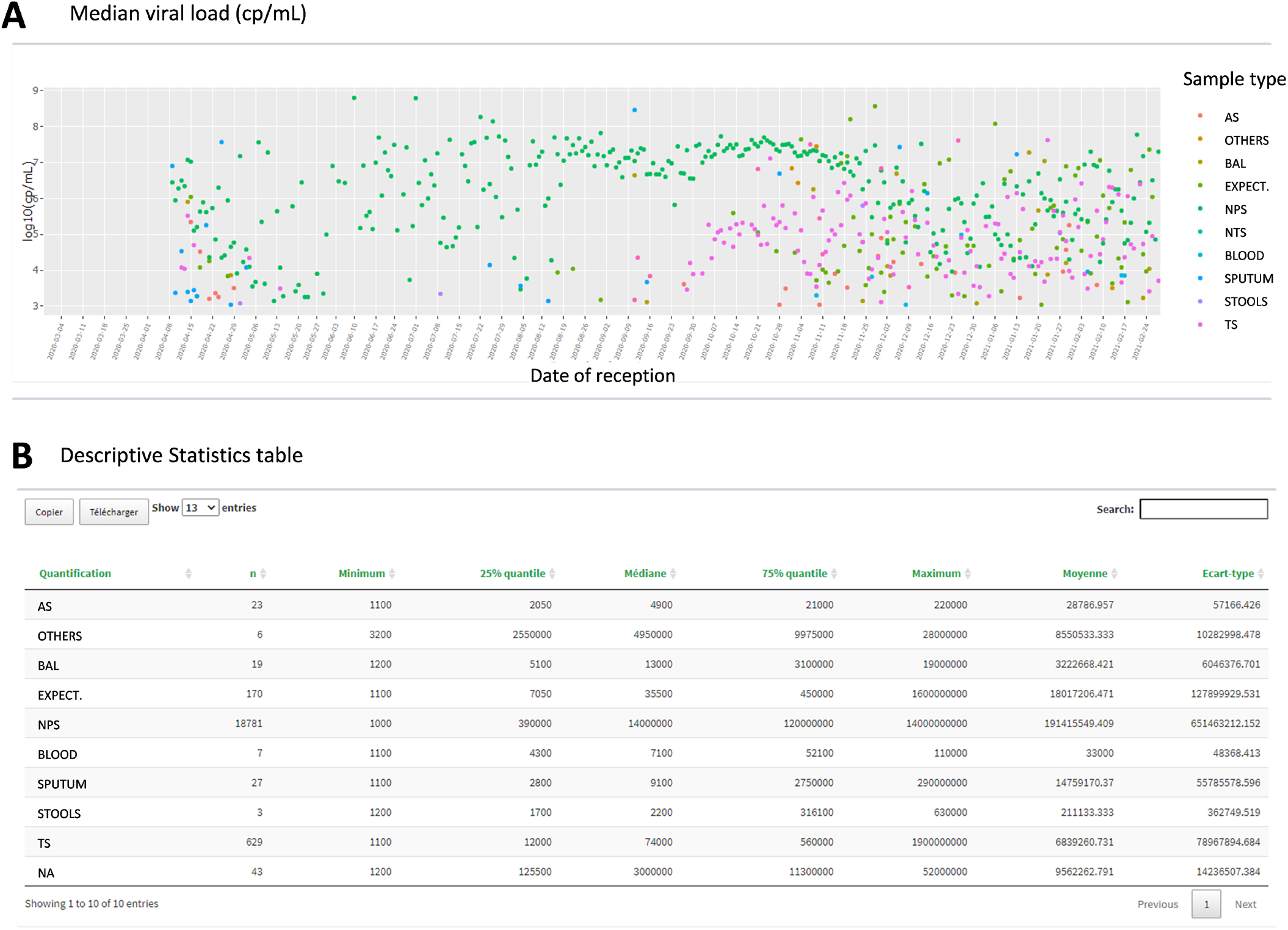
Screen shots of the Filter page. **A**. Viral load time series plot per sampling type, per date of reception and **B**. descriptive statistics table. AS: anal swab, BAL: bronchoalveolar lavage, EXPECT: expectoration; NPS, nasopharyngeal swab; TS: throat swab, NA: non applicable.

**Suppl. Figure 4:**
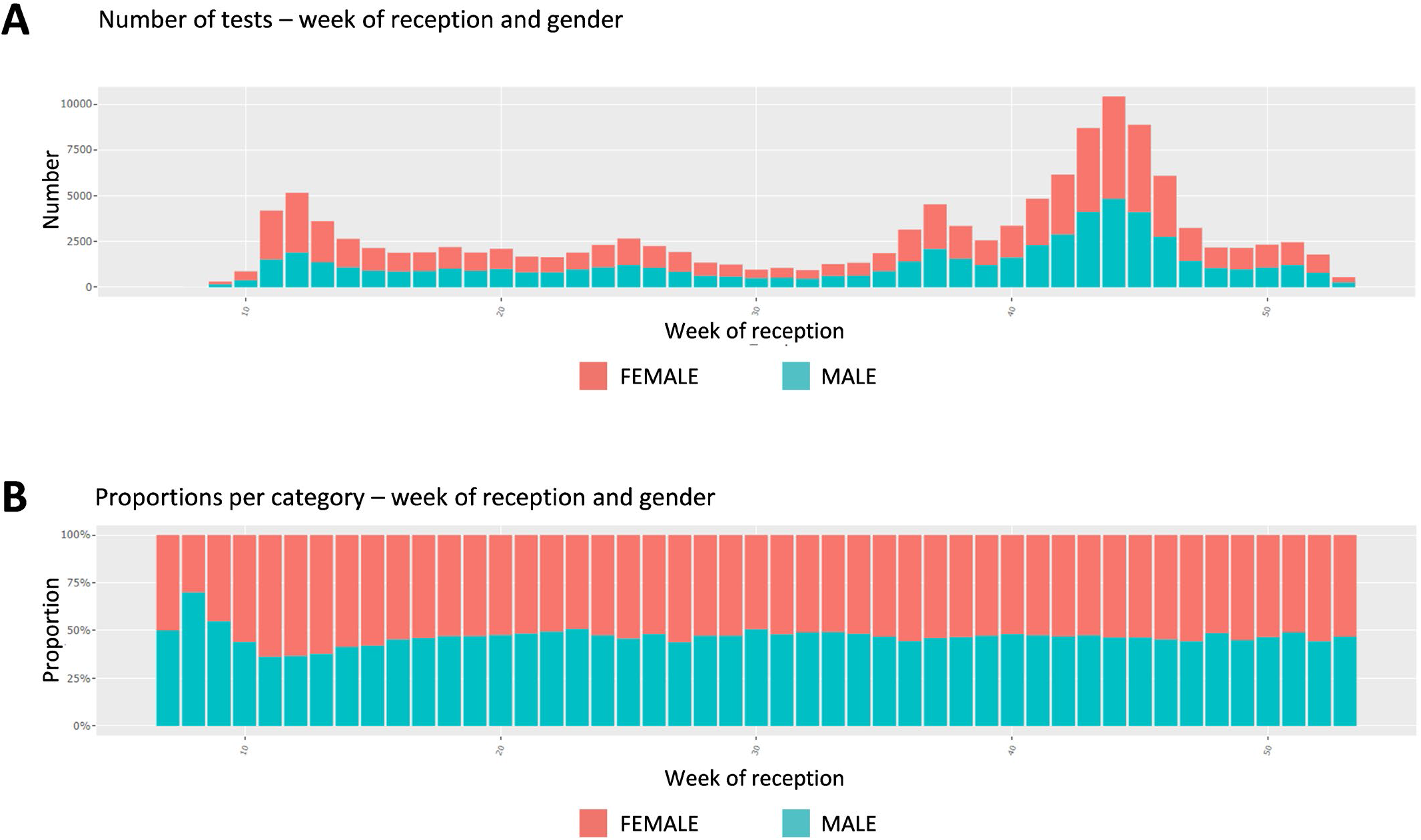
Screen shots of the Comparison page that allows 182 different combinations, focus on gender KPI. A. Number of tests per gender, per week of reception and B. proportion of each gender, per week of reception

